# Extent of *MGMT* promoter methylation modifies the effect of temozolomide on overall survival in patients with glioblastoma: a regional cohort study in Southeast Scotland

**DOI:** 10.1101/2021.07.22.21260997

**Authors:** Michael TC Poon, Shivank Keni, Vineeth Vimalan, Chak Ip, Colin Smith, Sara Erridge, Christopher J Weir, Paul M Brennan

**Author notes:** **Corresponding author** Dr Paul Brennan, Centre for Clinical Brain Sciences, University of Edinburgh, Edinburgh, BioQuarter, 49 Little France Crescent, Edinburgh, EH16 4SB. These authors contributed equally (Michael TC Poon, Shivank Keni). **Disclaimers** None.

## Abstract

**Background:** *MGMT* methylation in glioblastoma predicts response to temozolomide but dichotomising methylation status may mask the true prognostic value of quantitative *MGMT* methylation. This study evaluated whether extent of *MGMT* methylation interacts with the effect of temozolomide on overall survival.

**Methods:** We included consecutive glioblastoma patients diagnosed (April 2012-May 2020) at a neuro-oncology centre. All patients had quantitative *MGMT* methylation measured using pyrosequencing. Those with *MGMT* methylated tumours were stratified into high and low methylation groups based on a cut-off using Youden index on 2-year survival. Our accelerated failure time survival models included extent of *MGMT* methylation, age, post-operative Karnofsky performance score, extent of resection, temozolomide regimen and radiotherapy.

**Findings:** There were 414 patients. Optimal cut-off point using Youden index was 25.9% *MGMT* methylation. The number of patients in the unmethylated, low and high methylation groups was 223 (53.9%), 81 (19.6%) and 110 (26.6%), respectively. In the adjusted model, high (hazard ratio [HR] 0.60, 95% confidence intervals [CI] 0.46-0.79, p=0.005) and low (HR 0.67, 95%CI 0.50-0.89, p<0.001) methylation groups had better survival compared to unmethylated group. There was no evidence for interaction between *MGMT* methylation and completed temozolomide regimen (interaction term for low methylation p=0.097; high methylation p=0.071). This suggests no strong effect of *MGMT* status on survival in patients completing temozolomide regimen. In patients not completing the temozolomide regimen, higher *MGMT* methylation predicted better survival (interaction terms p<0.001).

**Interpretation:** Quantitative *MGMT* methylation may provide additional prognostic value. This is important when assessing clinical and research therapies.

## Introduction

O-6 methylguanine-DNA methyltransferase (*MGMT*) gene promoter methylation is an important biomarker in glioblastoma management. It is associated with better survival and is proposed to enhance efficacy of the alkylating agent temozolomide.^1–4^ Radiotherapy, with concurrent and adjuvant temozolomide, is the standard care ‘Stupp’ regimen for younger and fitter patients following glioblastoma diagnosis.

Measurements of methylation at the 5’-cytosine-phosphate-guanine-3’ (CpG) sites along the *MGMT* promoter determine the methylation status. Quantitative assays generate the percentage of methylated CpG sites, which is dichotomised against assay-specific thresholds.^5^ This dichotomy may mask additional prognostic values associated with extent of *MGMT* methylation.

Some studies have reported a positive association between extent of *MGMT* methylation and survival,^6–9^ but their smaller cohorts precluded analyses adjusting for confounders. They were also underpowered for examining effect modification of *MGMT* methylation on temozolomide, as predicted by the postulated mechanism of molecular interaction. This study assessed the association between extent of *MGMT* methylation and overall survival, and evaluated the interaction between *MGMT* methylation and temozolomide.

## Materials & Methods

### Study design & setting

This retrospective cohort study included consecutive patients reviewed by a regional neurooncology multidisciplinary team in Scotland, UK, between 1^st^ April 2012 and 31^st^ May 2020. Data collection was approved by Southeast Scotland Research Ethics Committee (reference 17/SS/0019).

### Participants

We included patients with histologically confirmed glioblastoma following surgery (biopsy or debulking). All patients had a first diagnosis of glioblastoma without prior low-grade glioma. Eligible patients were identified from weekly neuro-oncology multidisciplinary meetings. Data was collected from electronic patient records. Exclusion criteria included patients who: had incomplete treatment data; were lost to follow-up (>1 year since last review/treatment without documented follow-up and no evidence of death); had unavailable pre-operative neuroimaging; enrolled in a therapeutic clinical trial.

### Variables and data sources

Clinical characteristics included age at radiological diagnosis, sex, post-operative Karnofsky Performance Status (KPS), extent of resection, temozolomide administration, and *MGMT* methylation. Primary outcome was overall survival from date of radiological diagnosis. Secondary outcome was 2-year survival. Censoring date for survival data was 16^th^ January 2021.

Experienced board-certified neuropathologists made the histopathological diagnoses of glioblastoma. Isocitrate dehydrogenase 1 (*IDH1*) mutation testing using immunohistochemistry or sequencing was routine since 2017 and 2020, respectively. Routine magnetic resonance imaging (MRI) is performed within 72 hours post-operatively and extent of resection is reported by specialist neuroradiologists. Pyrosequencing using Qiagen *MGMT* assay generated the mean percentage methylation of the CpG sites (76-79) as the extent of *MGMT* methylation. The threshold for *MGMT* methylation was 6.4%, above which indicated the presence of *MGMT* methylation. We used the non-parametric method detailed in the Clinical and Laboratory Standards Institute guideline^10^ to generate the clinical threshold. Our institution offers patients radiotherapy with concurrent and adjuvant temozolomide as standard care.^11^ We recorded the number of weeks of concomitant and cycles of adjuvant temozolomide received. Temozolomide use was categorised into none, not completed (less than six weeks of concurrent or six adjuvant cycles) and standard ‘Stupp’ regimen (based on completed cycles). Whilst not specifically analysed in this study, patients who had previously received temozolomide within 12 months received procarbazine, lomustine and vincristine, or single agent lomustine on disease recurrence. Radiotherapy referred to the primary treatment received after glioblastoma diagnosis and was categorised into 60Gy, <60Gy (40Gy in 15 fractions or 30Gy in six fractions over two weeks), and none.

### Handling of variables

We reported categorical variables as frequencies with percentages and continuous variables as median values with quartiles and ranges. Post-operative KPS was categorised into <70 and ≥70 because of its relevance in informing treatment options. We classified the extent of resection into biopsy only, subtotal resection (<90% debulking), and gross total resection (≥90% debulking) of the contrast enhancing tumour, based on the residual volume identified in the standard care post-operative MRI. When considering the extent of *MGMT* methylation as a continuous variable, we applied a logarithmic transformation because of the positively skewed distribution. We performed stratification of *MGMT* methylation to assess whether there was a secondary threshold of prognostic value among patients with a methylated (>6.4%) glioblastoma. We constructed a receiver-operating characteristic (ROC) curve of logarithmically transformed *MGMT* methylation with 2-year survival as the dependent variable. We determined the optimal cut-off by maximising the Youden Index.

The resulting stratified *MGMT* methylation groups included unmethylated, low methylation and high methylation groups. Clinical *MGMT* status refers to the dichotomous *MGMT* status based on the clinical threshold (6.4%); logarithmically transformed *MGMT* level is the raw *MGMT* percentage methylation after natural logarithmic transformation.

### Bias and missing data

In the literature, pre-operative KPS is usually reported as a prognostic marker. However, oncologists use post-operative KPS as a marker of functional status to inform treatment selection. The inclusion of post-operative KPS made our analyses more robust in reflecting clinical decision making but not consistent with some studies. We did not have missing data to report because of our inclusion and exclusion criteria.

### Power calculation

The hazard ratio (HR) of death associated with *MGMT* methylation was 0.49 (95% confidence interval 0.41-0.59) in a systematic review and meta-analysis of clinical trial participants.^12^ We expected attenuation of this effect size outside the clinical trial setting. To estimate the power of our cohort, we set the hypothesised HR as 0.7, alpha at 5% and proportion of participants who died during follow-up as 95%. The calculated power was 94% with our sample size for analysis using a log-rank test. We anticipated the power of our analyses for detecting difference in *MGMT* methylation subgroups and for assessing interaction was lower.

### Statistical methods

We did not report p-values in descriptive tables to avoid multiple testing. We used the non-parametric Kaplan-Meier estimator to describe the survival function of *MGMT* methylation groups. We considered age, post-operative KPS, extent of resection, temozolomide use and radiotherapy as potential confounders based on the literature.^13^ The primary survival analysis of the association between *MGMT* methylation and overall survival was performed using the parametric accelerated failure time (AFT) model with Weibull distribution because of suspicion of hazard non-proportionality. Time ratios were converted to HRs for easier interpretation. We further performed these analyses in patients who had methylated (>6.4%) glioblastoma to assess the relevance in this subgroup, and in patients who received 60Gy of radiotherapy to assess confounding effect of radiotherapy. We also evaluated the effect of *MGMT* methylation in older patients aged 65 years or above.

For two-year mortality, we used multiple logistic regression adjusting for the same confounders other than radiotherapy and post-operative KPS because of complete data separation. Collinearity between variables in the multiple logistic regression was checked by examining the variance inflation factors.

To assess the effect modification between *MGMT* methylation and temozolomide use on survival, we introduced an interaction term between the two variables in the AFT model. Interaction was considered using the Wald test of the interaction term, and by comparing the interaction model with the main effects model using likelihood ratio test. We compared temozolomide cycles in patients receiving not-completed temozolomide regimen by *MGMT* methylation groups using Kruskal-Wallis test. We performed all data analyses in R (v4.1.0)^14^ using packages “cutpointr” (v1.1.0),^15^ “survival” (v3.2.7)^16^ and “survminer” (v0.4.8).^17^

## Results

### Participant characteristics

Of the 439 patients diagnosed with glioblastoma who met inclusion criteria, we excluded 25 patients due to: enrolment in a therapeutic clinical trial (N=7), lost to follow-up (N=15) and no pre-operative MRI scan (N=3). The final study cohort included 414 patients with total follow-up of 526 person-years (Table 1). The median age at diagnosis was 61 years (interquartile range [IQR]: 54-68 years). Post-operative KPS was 70 or above in 352 (85.0%) patients. Most patients (N=256; 61.8%) had debulking surgery and 158 (38.2%) had biopsy only. Sixty-six (15.9%) received the complete concomitant and adjuvant temozolomide Stupp regimen regardless of radiotherapy; 172 (41.5%) received fewer cycles. Most patients (59.9%) received 60Gy radiotherapy, whilst 28.5% patients received a lower radiotherapy dose (36 patients had 30Gy in 6 fractions over two weeks and 82 patients had 40Gy in 15 fractions over three weeks). Of the 226 patients who had *IDH1* mutation testing performed, 15 (6.6%) had an *IDH1-*mutant glioblastoma. Using the clinical threshold of 6.4% *MGMT* methylation, 191 (46.1%) patients had *MGMT* methylation. Characteristics of patients with unmethylated and methylated glioblastomas are presented in Supplementary Table 1.

**Table 1.**
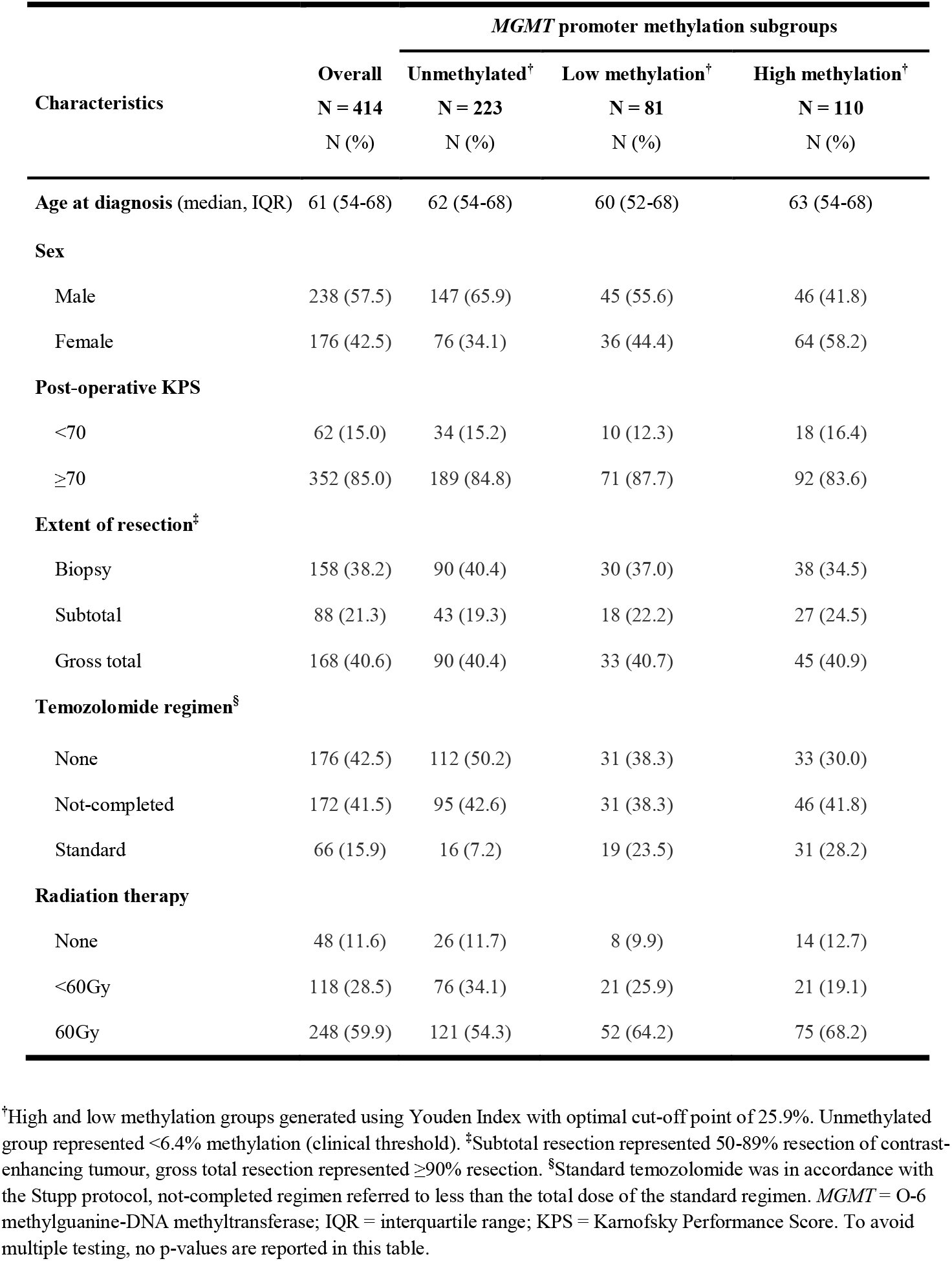
Demographic and clinical characteristics of 414 consecutive surgical patients with histopathologically confirmed glioblastoma in Southeast Scotland between April 2012 and May 2020

| Characteristics | *MGMT* promoter methylation subgroups |  |  |  |
| --- | --- | --- | --- | --- |
|  | Overall | Unmethylated<sup>†</sup> | Low methylation<sup>†</sup> | High methylation<sup>†</sup> |
|  | N = 414<br>N (%) | N = 223<br>N (%) | N = 81<br>N (%) | N = 110<br>N (%) |
| **Age at diagnosis** (median, IQR) | 61 (54-68) | 62 (54-68) | 60 (52-68) | 63 (54-68) |
| **Sex** |  |  |  |  |
| Male | 238 (57.5) | 147 (65.9) | 45 (55.6) | 46 (41.8) |
| Female | 176 (42.5) | 76 (34.1) | 36 (44.4) | 64 (58.2) |
| **Post-operative KPS** |  |  |  |  |
| <70 | 62 (15.0) | 34 (15.2) | 10 (12.3) | 18 (16.4) |
| ≥70 | 352 (85.0) | 189 (84.8) | 71 (87.7) | 92 (83.6) |
| **Extent of resection<sup>‡</sup>** |  |  |  |  |
| Biopsy | 158 (38.2) | 90 (40.4) | 30 (37.0) | 38 (34.5) |
| Subtotal | 88 (21.3) | 43 (19.3) | 18 (22.2) | 27 (24.5) |
| Gross total | 168 (40.6) | 90 (40.4) | 33 (40.7) | 45 (40.9) |
| **Temozolomide regimen<sup>§</sup>** |  |  |  |  |
| None | 176 (42.5) | 112 (50.2) | 31 (38.3) | 33 (30.0) |
| Not-completed | 172 (41.5) | 95 (42.6) | 31 (38.3) | 46 (41.8) |
| Standard | 66 (15.9) | 16 (7.2) | 19 (23.5) | 31 (28.2) |
| **Radiation therapy** |  |  |  |  |
| None | 48 (11.6) | 26 (11.7) | 8 (9.9) | 14 (12.7) |
| <60Gy | 118 (28.5) | 76 (34.1) | 21 (25.9) | 21 (19.1) |
| 60Gy | 248 (59.9) | 121 (54.3) | 52 (64.2) | 75 (68.2) |

<sup>†</sup>High and low methylation groups generated using Youden Index with optimal cut-off point of 25.9%. Unmethylated group represented <6.4% methylation (clinical threshold). <sup>‡</sup>Subtotal resection represented 50-89% resection of contrast-enhancing tumour, gross total resection represented ≥90% resection. <sup>§</sup>Standard temozolomide was in accordance with the Stupp protocol, not-completed regimen referred to less than the total dose of the standard regimen. *MGMT* = O-6 methylguanine-DNA methyltransferase; IQR = interquartile range; KPS = Karnofsky Performance Score. To avoid multiple testing, no p-values are reported in this table.

### MGMT promoter methylation

The median extent of *MGMT* methylation was 5.0% (IQR 3.0-27.8%). Among the 191 patients with methylated glioblastoma, the median *MGMT* methylation was 30.0% (IQR 16-42.8%). Figure 1 shows the distributions of *MGMT* methylation and its logarithmically transformed values. Based on the Youden Index, the optimal cut-off point of *MGMT* methylation for 2-year survival among patients with a methylated glioblastoma was 25.9% (3.25 in the logarithmic scale) with sensitivity and specificity of 67.8% and 46.6%, respectively. The resulting three groups comprised of unmethylated (N=223), low methylation (N=81) and high methylation (N=110) patients.

**Figure 1.**
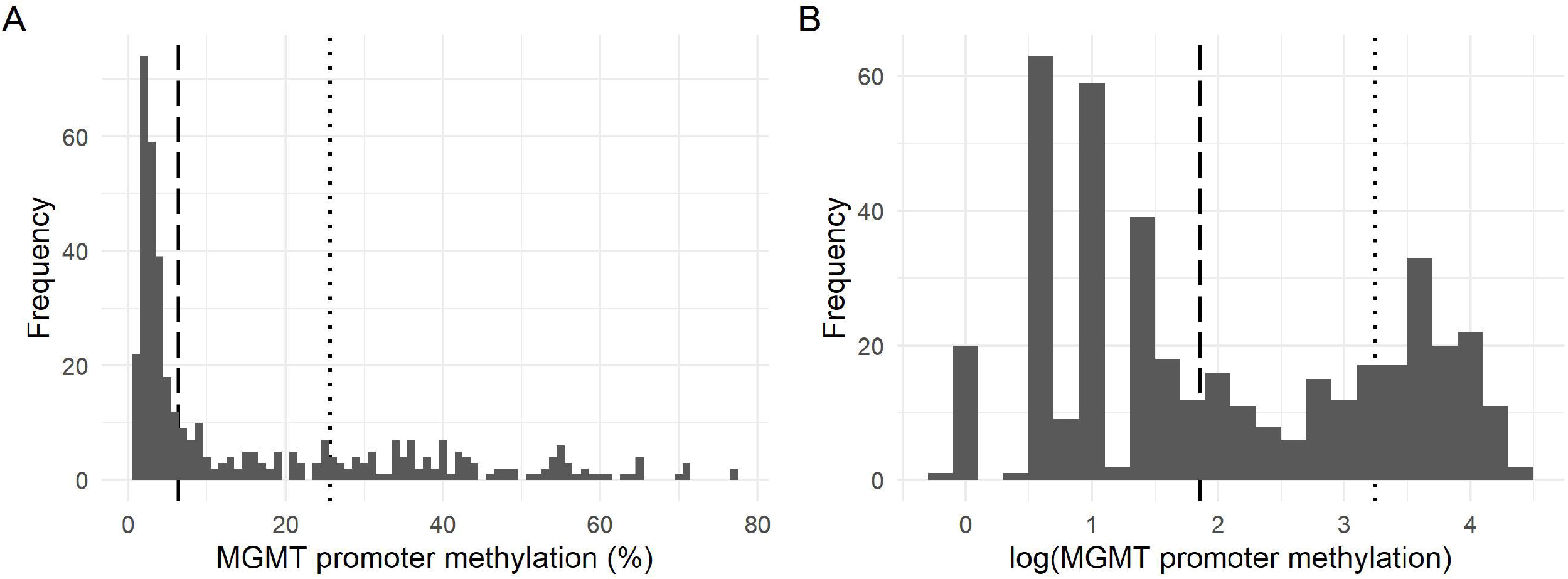
Histograms showing the extent of *MGMT* promoter methylation in the study cohort Vertical dashed line represents the clinical threshold of 6.4% in normal and logarithmic scale. Vertical dotted line represented the optimal cut-off point at 25.9% according to the Youden Index on 2-year survival. (A) Histogram of *MGMT* promoter methylation without transformation of data. (B) Histogram of logarithmically transformed *MGMT* promoter methylation level.

Characteristics of these groups are presented in Table 1. Higher proportions of patients in the low (23.5%) and high (28.2%) methylation groups received complete concomitant and adjuvant temozolomide compared to the unmethylated group (7.2%). Of the 15 patients with *IDH1* mutation, one patient had unmethylated, nine had low methylation, and five had high methylation glioblastoma.

### Overall survival and 2-year survival

There were 372 deaths observed during the follow-up period, of which 32 (8.6%) deaths occurred more than 2 years after tumour diagnosis. The median follow-up time was 0.90 years (IQR 0.51-1.56 years). Patients in the low and high *MGMT* methylation groups had longer median follow-up (1.21 and 1.07 years) compared to the unmethylated group (0.84 year). The proportions of patients surviving 2 years in the unmethylated, low and high methylation groups are 6.7% (n/N=15/223), 23.5% (n/N=19/81) and 36.4% (n/N=40/110), respectively. Figure 2 shows the survival functions of patients stratified by *MGMT* methylation groups. The median survival of patients who completed concomitant and adjuvant temozolomide cycles using Kaplan-Meier estimator was 24.7 months in unmethylated (N=14), 37.0 months in low methylation (N=19), and 36.3 months in high methylation groups (N=29).

**Figure 2.**
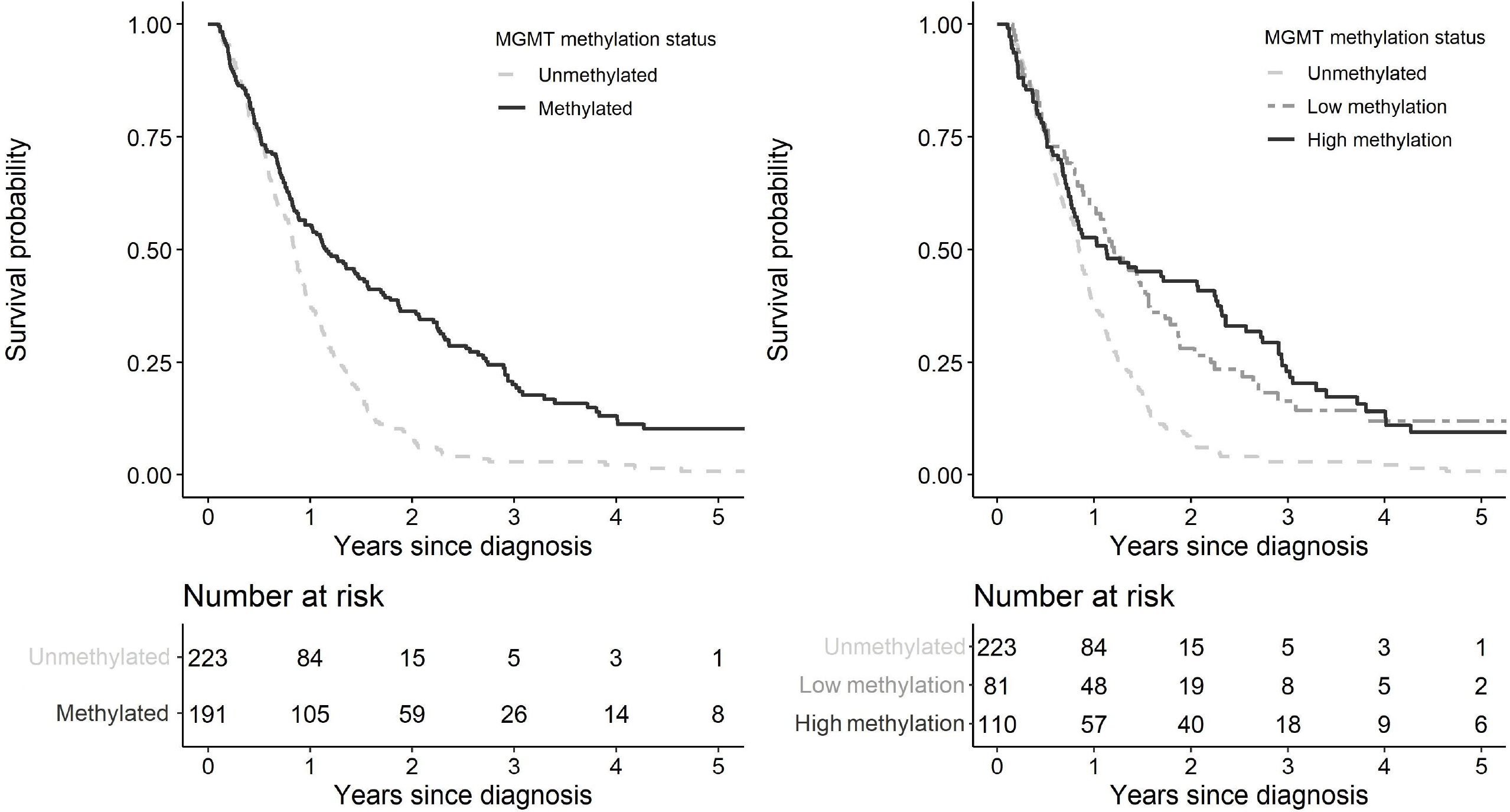
Kaplan-Meier curves of survival functions in 414 glioblastoma patients stratified by *MGMT* methylation status (A) Survival functions of patients with methylated and unmethylated glioblastoma based on clinical threshold (6.4%) of *MGMT* promoter methylation. (B) Survival functions of patients in the three methylation groups based on the clinical threshold and optimal cut-off point at 25.9%.

### Extent of MGMT methylation and overall survival

Considering the extent of *MGMT* methylation as a logarithmically transformed continuous variable, higher *MGMT* methylation was associated with better survival in both unadjusted (HR 0.73, 95%CI 0.67-0.79, p<0.001) and adjusted AFT models (HR 0.85, 95%CI 0.78-0.92, p<0.001) (Table 2).

**Table 2.** Multivariable survival analyses of 414 glioblastoma patients using accelerated failure time models

| Variables | No. of patients | Model 1 |  | Model 2 |  |
| --- | --- | --- | --- | --- | --- |
|  |  | HR (95% CI) | p-value | HR (95% CI) | p-value |
| ***MGMT* promoter methylation<sup>†</sup>** |  |  |  |  |  |
| Logarithmic continuous | 414 | 0.85 (0.78-0.92) | <0.001 | - | - |
| Unmethylated | 223 | - | - | ref | - |
| Low methylation | 81 | - | - | 0.67 (0.50-0.89) | 0.005 |
| High methylation | 110 | - | - | 0.60 (0.46-0.79) | <0.001 |
| **Temozolomide regimen<sup>§</sup>** |  |  |  |  |  |
| None | 176 | ref | - | ref | - |
| Not-completed | 172 | 0.49 (0.38-0.63) | <0.001 | 0.49 (0.38-0.63) | <0.001 |
| Standard | 66 | 0.17 (0.12-0.25) | <0.001 | 0.17 (0.12-0.26) | <0.001 |
| **Age at diagnosis (per year)** | 414 | 1.03 (1.02-1.04) | <0.001 | 1.03 (1.02-1.04) | <0.001 |
| **Post-operative KPS** |  |  |  |  |  |
| <70 | 62 | ref | - | ref | - |
| ≥70 | 352 | 0.46 (0.33-0.63) | <0.001 | 0.46 (0.33-1.63) | <0.001 |
| **Extent of Resection<sup>‡</sup>** |  |  |  |  |  |
| Biopsy | 158 | ref | - | ref | - |
| Subtotal | 88 | 0.92 (0.70-1.23) | 0.580 | 0.92 (0.69-1.23) | 0.578 |
| Gross total | 168 | 0.58 (0.45-0.75) | <0.001 | 0.58 (0.45-0.74) | <0.001 |
| **Radiation therapy** |  |  |  |  |  |
| None | 48 | ref | - | ref | - |
| <60Gy | 118 | 0.37 (0.26-0.53) | <0.001 | 0.36 (0.25-0.51) | <0.001 |
| 60Gy | 248 | 0.33 (0.22-0.49) | <0.001 | 0.32 (0.22-0.47) | <0.001 |

Model 1 used the logarithmically transformed *MGMT* methylation values; Model 2 used the three methylation groups determined by clinical threshold and the Youden Index. <sup>†</sup>High and low methylation groups generated using Youden Index with optimal cut-off point of 25.9%. Unmethylated group represented <6.4% methylation (clinical threshold).
<sup>‡</sup>Subtotal resection represented 50-89% resection of contrast-enhancing tumour, gross total resection represented ≥90% resection. <sup>§</sup>Standard temozolomide was in accordance with the Stupp protocol, not-completed regimen referred to less than the total dose of the standard regimen. HR = hazard ratio; CI = confidence interval; *MGMT* = O-6 methylguanine-DNA methyltransferase; KPS = Karnofsky Performance Score; ref = reference.

When restricting the analyses to 191 patients in the methylated groups, without stratifying into high and low extent of MGMT methylation, the logarithmically transformed *MGMT* methylation was not associated with overall survival (HR 0.89, 95%CI 0.71-1.12, p=0.314) (Supplementary Table 2). Using the methylation subgroups informed by the Youden Index, both low methylation (HR 0.67, 95%CI 0.50-0.89, p=0.005) and high methylation (HR 0.60, 95%CI 0.46-0.79, p<0.001) groups were associated with better survival compared with the unmethylated group (Table 2). Sensitivity analyses in 248 patients who received 60Gy of radiotherapy showed greater survival benefit associated with high methylation (HR 0.35, 95%CI 0.26-0.49, p<0.001) than with low methylation (HR 0.56, 95%CI 0.39-0.81, p=0.002) (Supplementary Table 3). In multiple logistic regression on 2-year mortality, *MGMT* methylation showed similar results (low methylation: odds ratio [OR] 0.30, 95%CI 0.12-0.76, p=0.011; high methylation: OR 0.13, 95%CI 0.05-0.29, p<0.001) (Supplementary Table 4).

### Extent of MGMT methylation in older patients

There were 168 patients aged ≥65 years at the time of glioblastoma diagnosis. Among the 76 (45.2%) patients with methylated glioblastoma, 46 had high methylation. In the multivariable logistic regression, *MGMT* methylation in either logarithmic scale (HR 0.87, 95%CI 0.76-1.00) or categories (low methylation: 0.64, 95%CI 0.41-1.01; high methylation: 0.70, 95%CI 0.45-1.09) was not associated with overall survival (Supplementary Table 5). Not-completed and standard temozolomide regimens were associated with better survival in the adjusted model; their corresponding HRs were 0.48 (95%CI 0.34-0.70, p<0.001) and 0.15 (95%CI 0.07-0.33, p<0.001), respectively.

### Effect modification of extent of MGMT promoter methylation by temozolomide regimen

Comparing models with and without interaction, likelihood ratio tests showed preference for the models with interaction terms between the extent of *MGMT* promoter methylation and temozolomide regimen (p<0.001). In the AFT model with additional interaction terms, logarithmically transformed *MGMT* methylation showed interaction with not-completed (interaction term p<0.001) and completed (p=0.028) temozolomide regimen (Table 3). Interaction between *MGMT* methylation and not-completed temozolomide regimen on overall survival was demonstrated when considering the three methylation groups (interaction terms p<0.001) (Table 3). The HRs associated with not-completed temozolomide regimen were 0.79 (95%CI 0.58-1.09, p=0.153) in unmethylated group, 0.29 (95%CI 0.18-0.45, p<0.001) in low methylation group and 0.27 (95%CI 0.18-0.40, p<0.001) in high methylation group. There was no evidence for interaction between *MGMT* methylation and standard temozolomide regimen (interaction term for low methylation group p=0.097; high methylation group p=0.071). The HRs associated with standard temozolomide regimen in all *MGMT* methylation groups were similar (Table 3). Supplementary Table 6 and 7 shows the adjusted HRs for other variables in the models. Sensitivity analysis of patients who received 60Gy of radiotherapy suggested a gradient across unmethylated (HR 0.77, 95 CI 0.50-1.19, p=0.235), low methylation (HR 0.36, 95%CI 0.19-0.68, p=0.002) and high methylation (0.21, 95%CI 0.12-0.35, p<0.001) groups in those receiving not-completed temozolomide (Table 3), though the smaller cohort precluded robust testing of an interaction.

**Table 3.** Interaction models between *MGMT* promoter methylation and temozolomide use on survival in 414 glioblastoma patients

|  | No. of patients | HR (95% CI) | p-value |
| --- | --- | --- | --- |
| **Model 1 – log(*MGMT*)** |  |  |  |
| Interaction between *MGMT* and TMZ<sup>§</sup> |  |  |  |
| No TMZ | - | ref | - |
| log(*MGMT*) + no TMZ | 176 | 1.07 (0.95-1.21) | 0.258 |
| log(*MGMT*) + not-completed TMZ regimen | 172 | 0.79 (0.55-1.13) | 0.197 |
| log(*MGMT*) + standard TMZ regimen | 66 | 0.25 (0.13-0.46) | <0.001 |
| **Model 2 – *MGMT* groups** |  |  |  |
| Interaction between *MGMT* and TMZ<sup>§</sup> |  |  |  |
| *No TMZ* |  |  |  |
| Unmethylated | 112 | ref | - |
| Low methylation | 31 | 1.20 (0.79-1.81) | 0.392 |
| High methylation | 33 | 1.18 (0.79-1.76) | 0.428 |
| *Not-completed TMZ* |  |  |  |
| Unmethylated | 95 | 0.79 (0.58-1.09) | 0.153 |
| Low methylation | 31 | 0.29 (0.18-0.45) | <0.001 |
| High methylation | 46 | 0.27 (0.18-0.40) | <0.001 |
| *Standard TMZ* |  |  |  |
| Unmethylated | 16 | 0.23 (0.13-0.42) | <0.001 |
| Low methylation | 19 | 0.13 (0.07-0.25) | <0.001 |
| High methylation | 31 | 0.13 (0.08-0.22) | <0.001 |
| **Model 3 – patients with 60Gy radiotherapy** |  |  |  |
| Interaction between *MGMT* and TMZ<sup>§</sup> |  |  |  |
| *No TMZ* |  |  |  |
| Unmethylated | 30 | ref | - |
| Low methylation | 13 | 0.81 (0.42-1.57) | 0.528 |
| High methylation | 10 | 0.50 (0.24-1.03) | 0.059 |
| *Not-completed TMZ* |  |  |  |
| Unmethylated | 77 | 0.77 (0.50-1.19) | 0.235 |
| Low methylation | 20 | 0.36 (0.19-0.68) | 0.002 |
| High methylation | 36 | 0.21 (0.12-0.35) | <0.001 |
| *Standard TMZ* |  |  |  |
| Unmethylated | 14 | 0.19 (0.09-0.38) | <0.001 |
| Low methylation | 19 | 0.10 (0.05-0.21) | <0.001 |
| High methylation | 29 | 0.09 (0.05-0.16) | <0.001 |

All models adjusted for age at diagnosis, post-operative Karnofsky Performance Score, extent of resection and radiotherapy. Hazard ratios were reparameterised from the models. Model 1 used the logarithmically transformed *MGMT* methylation values; Model 2 used the three methylation groups determined by clinical threshold and the Youden Index. <sup>§</sup>Standard temozolomide was in accordance with the Stupp protocol, not-completed regimen referred to less than the total dose of the standard regimen. Model 3 was the same as Model 2 but included patients who received 60Gy radiotherapy only. HR = hazard ratio; CI = confidence interval; *MGMT* = O-6 methylguanine-DNA methyltransferase; TMZ = temozolomide; ref = reference.

We examined whether the interaction with not-completed temozolomide regimen was due to differences in the number of temozolomide cycles received in each methylation group. Comparison between the groups did not demonstrate evidence of differences (p=0.092) (Supplementary Figure 1).

## Discussion

This retrospective cohort study of surgical patients diagnosed with glioblastoma in Scotland (UK) showed that *MGMT* promoter methylation was associated with better survival, but this effect was lost in patients aged ≥65 years old. *MGMT* promoter methylation modified the effects of temozolomide on survival for not-completed, but not standard temozolomide regimen, suggesting the interaction effect waned with increasing temozolomide up to the standard dose. This has implications for our interpretation of observational studies and clinical trials where *MGMT* status is only dichotomised and completion of standard temozolomide regimen is not considered. We need to clarify the presumptions about molecular mechanisms between *MGMT* methylation and temozolomide and whether the optimal number of temozolomide cycles vary between patients.

Our findings are consistent with previous studies that reported temozolomide is more beneficial in patients with *MGMT* methylated glioblastoma than in those with *MGMT* unmethylated glioblastoma.^18–21^ There is evidence supporting the biological mechanism of this observation. The MGMT protein removes alkyl groups from the O-6 guanine position, repairing the DNA damage caused by alkylating chemotherapies, such as temozolomide.^22^ Down-regulating *MGMT* expression by methylation of the *MGMT* promoter sequence therefore increases temozolomide sensitivity.^22,23^

Previous studies have reported an association between higher extent of *MGMT* promoter methylation and better overall survival.^6–8,24^ We observed longer survival in the higher methylation group compared to the low methylation group. Our larger cohort also demonstrated an interaction between the effect of multimodal therapy and *MGMT* methylation on overall survival. This contrasted a study using clinical trial data that reported no evidence of interaction, though the authors recognised the lack of power to test for effect modification.^18^ In our study, *MGMT* methylation modified the effect of temozolomide when the total concomitant and/or adjuvant dosage was less than the standard therapy. The absence of evidence for an interaction with standard temozolomide regimen may reflect the ceiling of benefit at this dose. This may have implications for our assumptions about the mechanism of interaction between temozolomide and *MGMT* methylation.

Two analyses have investigated the value of continuing temozolomide beyond adjuvant cycles after standard radiotherapy in glioblastoma.^25,26^ Both studies used a binary *MGMT* methylation status and reported no evidence of benefit from extended temozolomide in patients with methylated glioblastoma compared those with unmethylated glioblastoma (HR 0.89, 95%CI 0.63-1.26^25^ and HR 1.45, 95%CI 0.89-2.33).^26^ We observed similar benefit from standard temozolomide regimen across *MGMT* methylation groups (Table 3, Model 3). However, there may be a gradient effect (lower HRs with higher methylation) across methylation groups in those receiving not-completed temozolomide regimen. If this represents a biological interaction between *MGMT* methylation and temozolomide and our findings suggested a ceiling effect with standard temozolomide, the mix of patients with high and low methylation might have masked the benefit of extended temozolomide in previous studies. This warrants further interrogation.

Temozolomide administration in *MGMT* unmethylated patients remains controversial. Secondary analyses of the EORTC-NCIC trial data suggested minimal benefit for addition of temozolomide to radiotherapy in *MGMT* unmethylated patients.^18^ That study reported median survival of 11.8 months with radiotherapy only and 12.7 months with multimodal therapy. Withholding temozolomide based on *MGMT* methylation is debated because of limitations in methylation assays and lack of alternative therapies.^27^ We have shown that median survival for patients with unmethylated glioblastoma completing multimodal therapy was 26.1 months. The median survival for these patients was 17.1 months in another retrospective study.^7^ These observations may reflect careful patient selection for multimodal therapy in those with unmethylated tumours, although our analysis adjusted for factors affecting decision making such as age at diagnosis, functional status, and extent of resection. Standard multimodal therapy is still valuable for selected patients with *MGMT* unmethylated glioblastoma.

### *Strengths and* limitations

Our study measured quantitative *MGMT* methylation in consecutive glioblastoma patients over eight years, which allowed better power to examine the interaction between *MGMT* methylation and temozolomide. Because measuring *MGMT* methylation was routine, there was no missing data that would introduce bias. Residual confounding from other molecular markers and clinical variables could affect our results. However, we adjusted for the most relevant variables associated with survival and showed there was no differential distribution of *IDH1* mutation in the high methylation group. Selecting patients based on their clinical characteristics could have inflated benefits from multimodal therapy, particularly in the unmethylated group. We controlled for variables most influential to treatment decision and our cohort reflected routine clinical practice in contrast to selection bias associated with clinical trials. These made our study findings informative in using quantitative *MGMT* methylation in the clinical setting.

## Conclusion

Quantitative extent of *MGMT* promoter methylation has prognostic value in those receiving 60Gy radiotherapy. Patients receiving completed standard temozolomide regimen have comparable benefit regardless of *MGMT* methylation level. Assessing the extent of *MGMT* methylation in observational studies and clinical trials is necessary to permit accurate interpretation.

## Supporting information

STROBE checklist

## Data Availability

Please direct data request to the corresponding author.

