## Supplementary materials for "Extent of *MGMT* promoter methylation modifies the effect of temozolomide on overall survival in patients with glioblastoma: a regional cohort study in Southeast Scotland"

**Supplementary Table 1.** Demographic and clinical characteristics of 414 consecutive surgical patients with histopathologically confirmed glioblastoma in Southeast Scotland between April 2012 and May 2020 stratified by clinical threshold for *MGMT* promoter methylation

**Supplementary Table 2.** Multivariable survival analysis of 191 patients with methylated glioblastoma using an accelerated failure time model

**Supplementary Table 3**. Multivariable survival analysis of 248 patients with glioblastoma who received 60Gy of radiotherapy

**Supplementary Table 4**. Multiple logistic regression on deaths within 2 years of glioblastoma diagnosis

**Supplementary Table 5**. Multivariable survival analysis of 138 older (≥65 years) patients

**Supplementary Table 6**. Interaction models between logarithmically transformed *MGMT* promoter methylation and temozolomide use on survival in 414 glioblastoma patients

**Supplementary Table 7**. Interaction models between *MGMT* promoter methylation subgroups and temozolomide use on survival in 414 glioblastoma patients

**Supplementary Figure 1.** Histograms showing the distribution of total concomitant and adjuvant temozolomide cycles stratified by *MGMT* methylation groups

**Supplementary Table 1.** Demographic and clinical characteristics of 415 consecutive surgical patients with histopathologically confirmed glioblastoma in Southeast Scotland between April 2012 and May 2020 stratified by clinical threshold for *MGMT* promoter methylation

| **Characteristics** | **Overall**  **N = 414**  N (%) | **Unmethylated^†^**  **N = 223**  N (%) | **Methylated^†^**  **N = 191**  N (%) |
| --- | --- | --- | --- |
| **Age at diagnosis** (median, IQR) | 61 (54-68) | 62 (54-68) | 61 (53-68) |
| **Sex** |  |  |  |
| Male | 238 (57.5) | 147 (65.9) | 91 (47.6) |
| Female | 176 (42.5) | 76 (34.1) | 100 (52.4) |
| **Post-operative KPS** |  |  |  |
| <70 | 62 (15.0) | 34 (15.2) | 28 (14.7) |
| 70+ | 352 (85.0) | 189 (84.8) | 163 (85.3) |
| **Extent of resection^‡^** |  |  |  |
| Biopsy | 158 (38.2) | 90 (40.4) | 68 (35.6) |
| Subtotal | 88 (21.3) | 43 (19.3) | 45 (23.6) |
| Gross total | 168 (40.6) | 90 (40.4) | 78 (40.8) |
| **Temozolomide regimen^§^** |  |  |  |
| None | 176 (42.5) | 112 (50.2) | 64 (33.5) |
| Non-completed | 172 (41.5) | 95 (42.6) | 77 (40.3) |
| Standard | 66 (15.9) | 16 (7.2) | 50 (26.2) |
| **Radiotherapy** |  |  |  |
| None | 48 (11.6) | 26 (11.7) | 22 (11.5) |
| <60Gy | 118 (28.5) | 76 (34.1) | 42 (22.0) |
| 60Gy | 248 (59.9) | 121 (54.3) | 127 (66.5) |

**^†^**Clinical threshold for *MGMT* promoter methylation was 6.4%, above which indicated the presence of *MGMT* methylation. **^‡^**Subtotal resection represented 50-89% resection of contrast-enhancing tumour, gross total resection represented ≥90% resection. **^§^**Standard temozolomide was in accordance with the Stupp protocol, non-completed regimen referred to less than the total dose of the standard regimen. IQR = interquartile range; KPS = Karnofsky Performance Score. To avoid multiple testing, no p-values are reported in this table.

**Supplementary Table 2.** Multivariable survival analysis of 191 patients with methylated glioblastoma using an accelerated failure time model

| **Variable** | **No. of patients** | **HR (95% CI)** | **p-value** |
| --- | --- | --- | --- |
| ***MGMT* promoter methylation** |  |  |  |
| Logarithmic continuous | 191 | 0.89 (0.71-1.12) | 0.314 |
| **Temozolomide regimen^§^** |  |  |  |
| None | 64 | ref | - |
| Non-completed | 77 | 0.31 (0.21-0.47) | <0.001 |
| Standard | 50 | 0.16 (0.10-0.27) | <0.001 |
| **Age at diagnosis (per year)** | 191 | 1.03 (1.01-1.04) | 0.002 |
| **Post-operative KPS** |  |  |  |
| <70 | 28 | ref | - |
| ≥70 | 163 | 0.33 (0.20-1.55) | 0.440 |
| **Extent of Resection^‡^** |  |  |  |
| Biopsy | 68 | ref | - |
| Subtotal | 45 | 0.97 (0.61-1.55) | 0.896 |
| Gross total | 78 | 0.49 (0.33-0.75) | <0.001 |
| **Radiotherapy** |  |  |  |
| None | 22 | ref | - |
| <60Gy | 42 | 0.26 (0.14-0.48) | <0.001 |
| 60Gy | 127 | 0.22 (0.12-0.40) | <0.001 |

**^‡^**Subtotal resection represented 50-89% resection of contrast-enhancing tumour, gross total resection represented ≥90% resection. **^§^**Standard temozolomide was in accordance with the Stupp protocol, non-completed regimen referred to less than the total dose of the standard regimen. HR = Hazard ratio; CI = confidence interval; KPS = Karnofsky Performance Score.

**Supplementary Table 3**. Multivariable survival analysis of 248 patients with glioblastoma who received 60Gy of radiotherapy

| **Variables** | **No. of patients** | **Model 1** | | **Model 2** | |
| --- | --- | --- | --- | --- | --- |
|  |  | **HR (95% CI)** | **p-value** | **HR (95% CI)** | **p-value** |
| ***MGMT* promoter methylation^†^** |  |  |  |  |  |
| Logarithmic continuous | 248 | 0.70 (0.63-0.78) | **<**0.001 | - | - |
| Unmethylated | 121 | - | - | ref | - |
| Low methylation | 52 | - | - | 0.56 (0.39-0.81) | 0.002 |
| High methylation | 75 | - | - | 0.35 (0.26-0.49) | <0.001 |
| **Temozolomide regimen^§^** |  |  |  |  |  |
| None | 53 | ref | - | ref | - |
| Non-completed | 133 | 0.63 (0.45-0.90) | 0.011 | 0.62 (0.44-0.88) | 0.007 |
| Standard | 62 | 0.20 (0.13-0.31) | <0.001 | 0.19 (0.12-0.30) | <0.001 |
| **Age at diagnosis (per year)** | 248 | 1.04 (1.02-1.05) | <0.001 | 1.04 (1.02-1.05) | <0.001 |
| **Post-operative KPS** |  |  |  |  |  |
| <70 | 9 | ref | - | ref | - |
| ≥70 | 239 | 0.21 (0.10-0.41) | <0.001 | 0.20 (0.10-0.41) | <0.001 |
| **Extent of Resection^‡^** |  |  |  |  |  |
| Biopsy | 58 | ref | - | ref | - |
| Subtotal | 57 | 1.40 (0.94 -2.08) | 0.098 | 1.34 (0.90-2.00) | 0.148 |
| Gross total | 133 | 0.66 (0.47-0.94) | 0.019 | 0.68 (0.48-0.96) | 0.029 |

Model 1 used the logarithmically transformed *MGMT* methylation values; Model 2 used the three methylation groups determined by clinical threshold and the Youden Index. **^†^**High and low methylation groups generated using Youden Index with optimal cut-off point of 25.9%. Unmethylated group represented <6.4% methylation (clinical threshold). **^‡^**Subtotal resection represented 50-89% resection of contrast-enhancing tumour, gross total resection represented ≥90% resection. **^§^**Standard temozolomide was in accordance with the Stupp protocol, non-completed regimen referred to less than the total dose of the standard regimen. HR = hazard ratio; CI = confidence interval; *MGMT* = O-6 methylguanine-DNA methyltransferase; KPS = Karnofsky Performance Score; ref = reference.

**Supplementary Table 4**. Multiple logistic regression on deaths within 2 years of glioblastoma diagnosis

| **Variables** | **No. of patients** | **Model 1** | | **Model 2** | |
| --- | --- | --- | --- | --- | --- |
|  |  | **OR (95% CI)** | **p-value** | **OR (95% CI)** | **p-value** |
| ***MGMT* promoter methylation^†^** |  |  |  |  |  |
| Logarithmic continuous | 414 | 0.45 (0.33-0.61) | <0.001 | - | - |
| Unmethylated | 223 | - | - | ref | - |
| Low methylation | 81 | - | - | 0.33 (0.13-0.84) | 0.019 |
| High methylation | 110 | - | - | 0.14 (0.06-0.32) | <0.001 |
| **Temozolomide regimen^§^** |  |  |  |  |  |
| None | 176 | ref | - | ref | - |
| Non-completed | 172 | 0.16 (0.04-0.52) | 0.006 | 0.15 (0.03-0.49) | 0.004 |
| Standard | 66 | 0.02 (0.00-0.07) | <0.001 | 0.02 (0.00-0.09) | <0.001 |
| **Age at diagnosis (per year)** | 414 | 1.05 (1.02-1.08) | <0.001 | 1.05 (1.02-1.08) | <0.001 |
| **Extent of Resection^‡^** |  |  |  |  |  |
| Biopsy | 158 | ref | - | ref | - |
| Subtotal | 88 | 0.99 (0.34-2.94) | 0.986 | 0.86 (0.30-2.50) | 0.800 |
| Gross total | 168 | 0.41 (0.16-0.98) | 0.049 | 0.42 (0.17-0.99) | 0.053 |

Model 1 used the logarithmically transformed *MGMT* methylation values; Model 2 used the three methylation groups determined by clinical threshold and the Youden Index. **^†^**High and low methylation groups generated using Youden Index with optimal cut-off point of 25.9%. Unmethylated group represented <6.4% methylation (clinical threshold). **^‡^**Subtotal resection represented 50-89% resection of contrast-enhancing tumour, gross total resection represented ≥90% resection. **^§^**Standard temozolomide was in accordance with the Stupp protocol, non-completed regimen referred to less than the total dose of the standard regimen. OR = Odds ratio; CI = confidence interval; *MGMT* = O-6 methylguanine-DNA methyltransferase; KPS = Karnofsky Performance Score; ref = reference.

**Supplementary Table 5**. Multivariable survival analysis of 168 older (≥65 years) patients

| **Variables** | **No. of patients** | **Model 1** | | **Model 2** | |
| --- | --- | --- | --- | --- | --- |
|  |  | **HR (95% CI)** | **p-value** | **HR (95% CI)** | **p-value** |
| ***MGMT* promoter methylation^†^** |  |  |  |  |  |
| Logarithmic continuous | 168 | 0.86 (0.75-0.99) | 0.03 | - | - |
| Unmethylated | 92 | - | - | ref | - |
| Low methylation | 30 | - | - | 0.60 (0.39-0.94) | 0.026 |
| High methylation | 46 | - | - | 0.67 (0.43-1.05) | 0.0.81 |
| **Temozolomide regimen^§^** |  |  |  |  |  |
| None | 93 | ref | - | ref | - |
| Non-completed | 65 | 0.48 (0.33-0.69) | <0.001 | 0.48 (0.34-0.70) | <0.001 |
| Standard | 10 | 0.15 (0.07-0.34) | <0.001 | 0.15 (0.07-0.33) | <0.001 |
| **Age at diagnosis (per year)** | 168 | 0.94 (0.89-1.00) | 0.042 | 0.95 (0.90-1.00) | 0.056 |
| **Post-operative KPS** |  |  |  |  |  |
| <70 | 25 | ref | - | ref | - |
| ≥70 | 143 | 0.34 (0.21-0.55) | <0.001 | 0.34 (0.21-0.55) | <0.001 |
| **Extent of Resection^‡^** |  |  |  |  |  |
| Biopsy | 63 | ref | - | ref | - |
| Subtotal | 40 | 0.84 (0.52-1.36) | 0.475 | 0.82 (0.51-1.33) | 0.425 |
| Gross total | 65 | 0.44 (0.28-0.68) | <0.001 | 0.41 (0.27-0.64) | <0.001 |
| **Radiotherapy** |  |  |  |  |  |
| None | 23 | ref | - | ref | - |
| <60Gy | 82 | 0.58 (0.35-0.97) | 0.037 | 0.57 (0.34-0.95) | 0.031 |
| 60Gy | 63 | 0.40 (0.22-0.76) | 0.005 | 0.39 (0.21-0.74) | 0.004 |

Model 1 used the logarithmically transformed *MGMT* methylation values; Model 2 used the three methylation groups determined by clinical threshold and the Youden Index. **^†^**High and low methylation groups generated using Youden Index with optimal cut-off point of 25.9%. Unmethylated group represented <6.4% methylation (clinical threshold). **^‡^**Subtotal resection represented 50-89% resection of contrast-enhancing tumour, gross total resection represented ≥90% resection. **^§^**Standard temozolomide was in accordance with the Stupp protocol, non-completed regimen referred to less than the total dose of the standard regimen. HR = hazard ratio; CI = confidence interval; *MGMT* = O-6 methylguanine-DNA methyltransferase; KPS = Karnofsky Performance Score; ref = reference.

**Supplementary Table 6**. Interaction models between logarithmically transformed *MGMT* promoter methylation and temozolomide use on survival in 414 glioblastoma patients

|  | **HR (95% CI)** | **p-value** |
| --- | --- | --- |
| **Interaction between *MGMT* and TMZ^§^** |  |  |
| No TMZ | ref | - |
| log(*MGMT*) + no TMZ | 1.07 (0.95-1.21) | 0.258 |
| log(*MGMT*) + non-completed TMZ regimen | 0.79 (0.55-1.13) | 0.197 |
| log(*MGMT*) + standard TMZ regimen | 0.25 (0.13-0.46) | <0.001 |
| **Age at diagnosis (per year)** | 1.03 (1.01-1.04) | <0.001 |
| **Post-operative KPS** |  |  |
| <70 | ref | - |
| ≥70 | 0.41 (0.30-0.56) | <0.001 |
| **Extent of Resection^‡^** |  |  |
| Biopsy | ref | - |
| Subtotal | 0.99 (0.75-1.32) | 0.969 |
| Gross total | 0.55 (0.43-0.71) | <0.001 |
| **Radiotherapy** |  |  |
| None | ref | - |
| <60Gy | 0.35 (0.24-0.49) | <0.001 |
| 60Gy | 0.29 (0.19-0.42) | <0.001 |

Hazard ratios were reparameterised from the models. Model 1 used the logarithmically transformed *MGMT* methylation values; Model 2 used the three methylation groups determined by clinical threshold and the Youden Index. **^§^**Standard temozolomide was in accordance with the Stupp protocol, non-completed regimen referred to less than the total dose of the standard regimen. HR = hazard ratio; CI = confidence interval; *MGMT* = O-6 methylguanine-DNA methyltransferase; TMZ = temozolomide; ref = reference.

**Supplementary Table 7**. Interaction models between *MGMT* promoter methylation subgroups and temozolomide use on survival in 414 glioblastoma patients

|  | **HR (95% CI)** | **p-value** |
| --- | --- | --- |
| **Interaction between *MGMT* and TMZ^§^** |  |  |
| Unmethylated + no TMZ | ref | - |
| Unmethylated + non-completed TMZ regimen | 0.79 (0.58-1.09) | 0.153 |
| Unmethylated + standard TMZ regimen | 0.23 (0.13-0.42) | <0.001 |
| Low methylation + no TMZ | 1.20 (0.79-1.81) | 0.392 |
| Low methylation + non-completed TMZ regimen | 0.29 (0.18-0.45) | <0.001 |
| Low methylation + standard TMZ regimen | 0.13 (0.07-0.25) | <0.001 |
| High methylation + no TMZ | 1.18 (0.79-1.76) | 0.428 |
| High methylation + non-completed TMZ regimen | 0.27 (0.18-0.40) | <0.001 |
| High methylation + standard TMZ regimen | 0.13 (0.08-0.22) | <0.001 |
| **Age at diagnosis (per year)** | 1.02 (1.01-1.03) | <0.001 |
| **Post-operative KPS** |  |  |
| <70 | ref | - |
| ≥70 | 0.41 (0.30-0.57) | <0.001 |
| **Extent of Resection^‡^** |  |  |
| Biopsy | ref | - |
| Subtotal | 0.97 (0.73-1.29) | 0.824 |
| Gross total | 0.54 (0.42-0.70) | <0.001 |
| **Radiotherapy** |  |  |
| None | ref | - |
| <60Gy | 0.37 (0.26-0.53) | <0.001 |
| 60Gy | 0.29 (0.20-0.43) | <0.001 |

Hazard ratios were reparameterised from the models. Model 1 used the logarithmically transformed *MGMT* methylation values; Model 2 used the three methylation groups determined by clinical threshold and the Youden Index. **^§^**Standard temozolomide was in accordance with the Stupp protocol, non-completed regimen referred to less than the total dose of the standard regimen. HR = hazard ratio; CI = confidence interval; *MGMT* = O-6 methylguanine-DNA methyltransferase; TMZ = temozolomide; ref = reference.

**Supplementary Figure 1.** Histograms showing the distribution of total concomitant and/or adjuvant temozolomide cycles stratified by *MGMT* methylation groups in those receiving non-completed temozolomide regimen

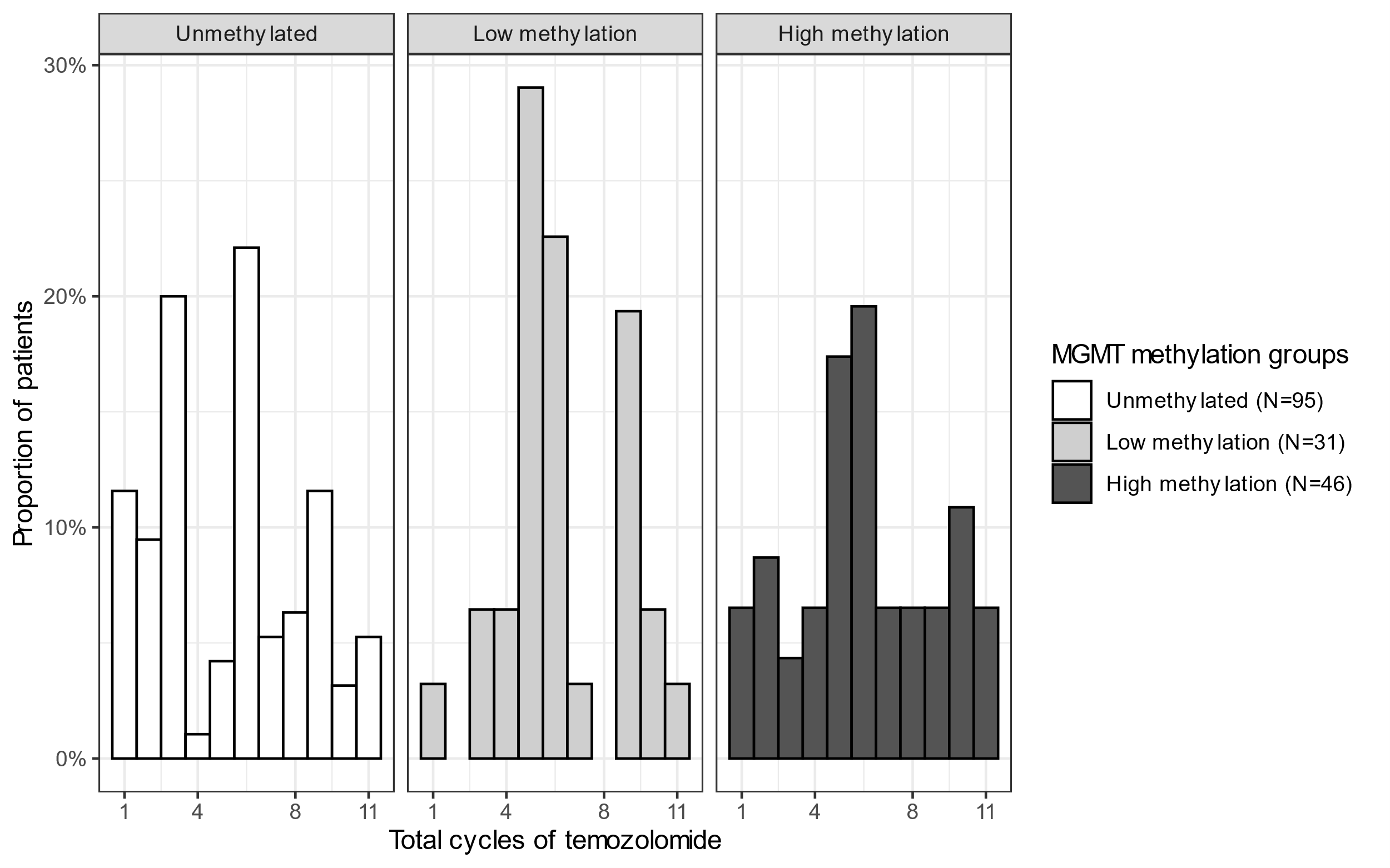

There were 172 patients receiving less than standard temozolomide regimen. 6 weeks of concomitant daily temozolomide with radiotherapy were considered 6 cycles. Adjuvant temozolomide cycles referred to 28-day cycles when temozolomide was taken for 5 days within each cycle.
